# Perceptions of immunity and vaccination certificates among the general population in Geneva, Switzerland

**DOI:** 10.1101/2021.06.22.21259189

**Authors:** Mayssam Nehme, Helene Baysson, Nick Pullen, Ania Wisniak, Francesco Pennacchio, MarÍa-Eugenia Zaballa, Vanessa Fargnoli, Didier Trono, Laurent Kaiser, Samia Hurst, Claudine Burton-Jeangros, Silvia Stringhini, Specchio-COVID19 study group, Idris Guessous

## Abstract

As of June 2021, the European Union (EU) and Switzerland have published information about the introduction of COVID certificates in order to facilitate the safe free movement of their citizens. With implementation underway, little is known about the public perception of such certificates with potential differences in acceptability among individuals.

In March 2021, a self-administered online questionnaire was proposed to all individuals 18 years and older participating in the longitudinal follow-up of population-based seroprevalence studies in Geneva, Switzerland. The questionnaire covered aspects of individual and collective benefits, while allowing participants to select contexts in which vaccination certificates should be presented. Results were presented as the proportion of individuals agreeing or disagreeing with the implementation of vaccination certificates, selecting specific contexts where certificates should be presented, and agreeing or disagreeing with the potential risks related to certificates. Logistic regression was used to calculate odds ratios for factors associated with certificate non-acceptance.

Overall, 4,056 individuals completed the questionnaire (response rate 77.6%; mean age 53.3 ± standard deviation 14.4 years; 56.1% were women). About 61.0% of participants agreed or strongly agreed that a vaccination certificate was necessary in certain contexts; and 21.6% believed there was no context where vaccination certificates should be presented. Contexts where a majority of participants perceived a vaccination certificate should be presented included jobs where others would be at risk of COVID-related complications (60.7%), jobs where employees would be at risk of getting infected (58.7%), or to be exempt from quarantine when traveling abroad (56.1%). Contexts where fewer individuals perceived the need for vaccination certificates to be presented were participation in large gatherings (36.9%), access to social venues (35.5%), or sharing the same workspace (21.5%). Younger age, an absence of willingness to get vaccinated, and an absence of belief in vaccination as an important step in surmounting the pandemic were factors associated with certificate non-acceptance.

This large population-based study showed that the general adult population in Geneva, Switzerland, agreed with the implementation of vaccination certificates in work-related and travel-related contexts. However, this solution was perceived as unnecessary for access to large gatherings or social venues, or to share the same workspace. Differences were seen with gender, age, education, socio-economic status, and vaccination willingness and perception, highlighting the importance of taking personal and sociodemographic variations into consideration when predicting acceptance of such certificates.

## Introduction

The COVID-19 pandemic will continue to have an impact on several dimensions of physical and mental health, as well as on social and economic parameters for years to come^1,2,3,4^. With the advent of effective vaccines, mass vaccination is recognized as an effective way out of the pandemic, especially taking into account that any public health restrictive measures should be in adequate response to specific and demonstrable risk^5^. Countries with extensive vaccination programs have already implemented “green passes”^6,7^, and the European Union is entering the deployment phase of its recently conceived COVID certificates^8^, in an effort to resume and once again allow free movement^9^. Switzerland has announced a COVID certificate will be available by June 2021^10^. COVID certificates could attest to an individual’s vaccination status, a past SARS-CoV-2 infection or the absence of current infection^9^. As of June 2021, COVID certificates in Switzerland will be used for international travel, large gatherings of at least 1’000 individuals and at clubs, discos and dance events^11^.

While implementation is underway, little is known about the public perception of COVID certificates. There has been conflicting evidence about the role of COVID certificates in vaccination programs uptake, and as a strategy in a phased reduction of lockdown measures^12,13^. Vaccination certificates could allow a safer access to several activities, and may increase the uptake of immunization when considering an incentive-based approach^14,15^. However, they could also be viewed as coercive measures creating a backlash and further increasing any pre-existing resistance to vaccination^14,16^. A recent review of the public perception of COVID certificates, and their potential impact on behavior, and the uptake of testing and vaccination reported different acceptance rates depending on context (travel, social or professional)^17^. There was little information on sociodemographic differences in most of the studies included in this review^17^. A survey addressed to 12,000 scientists revealed their overall favorable attitudes towards COVID certificates^18^. Scientists perceived immunity certificates favorably for their positive impact on public health and the economy, despite highlighting risks related to equity and equality of the implementation process. Differences were perceived among participants as US-based scholars, men and scientists with more conservative political views were overall more favorable to immunity certificates ^18^.

To date, there is little information about the general population-based acceptance and perception of COVID certificates. In November 2020^19^, we published results on the perception of immunity certificates mostly in relation to natural immunity. We now present updated results (March 2021) which include population perception on vaccination certificates after it became clear that mass vaccination would become available, with a larger sample size of participants as well as additional stratifications.

## Methods

### Study setting and data collection

In March 2021, a self-administered online questionnaire was proposed to all individuals 18 years and older participating in the longitudinal follow-up of population-based seroprevalence studies in Geneva, Switzerland^20,21,22^. This longitudinal follow-up is conducted via the Specchio-COVID19 digital platform allowing participants to answer regular online questionnaires^23^.

All individuals gave consent, and the study was approved by the Cantonal Research Ethics Commission of Geneva, Switzerland (protocol numbers CER 2020-01540 and CER 2020-00881). Questions about vaccination certificates were part of a larger vaccination questionnaire. These specific questions were elaborated based on the results of the initial questionnaire on the perception of immunity certificates^19^, as well as the results of a qualitative study conducted between July and November 2020, aiming at identifying arguments for or against immunity certificates^24^. An initial invitation to complete the questionnaire was sent by e-mail on March 17, 2021 with a reminder two weeks later.

The questionnaire (supplementary table S1) was collaboratively constructed by physicians (IG, MN), epidemiologists (IG, SS, HB), sociologists (CBJ, VF), and an ethicist (SH). Two main questions were asked: 1-Select the context(s) in which a vaccination certificate should be presented (with a list of contexts); 2-What is your opinion about the following statements on the implementation of a COVID-19 vaccination certificate (with a list of statements)? The answers to the latter question were based on a 5-point Likert scale with the following categories: 1 “Strongly disagree”; 2 “Disagree”; 3 “Neither agree nor disagree”; 4” Agree”; 5” Strongly agree” (Supplemental table S1 for details).

### Analysis

We used the statistical software STATA version 15.1. Descriptive analyses included percentages with comparisons using chi-square tests. P-values were considered significant at p<0.05. Stratifications were based on age categories, gender, education level, household income, employment status, occupational position, past SARS-CoV-2 infection, vaccination willingness, and vaccination perception.

Household income was calculated taking into consideration household revenue and the number of individuals in a household. Household income was then compared to the cantonal database available online^25^ with the categories defined as “Low” (Below first quartile); “Mid” (Between quartiles 1 and 3); “High” (Higher than the third quartile). Education was categorized as follows: “Lowest level” included compulsory education and no formal education; “Lower level” included apprenticeships; “Higher level” included secondary and specialized schools; “Highest level” included universities, higher professional education, and doctorates. Occupational position was categorized as follows: Blue collar workers were qualified employees practicing manual labor, craftsmen, traders, farmers and employees without specific training; Lower grade white collar workers were qualified employees (non-manual labor); Higher grade white collar workers were employees with a profession requiring intermediate training; Professional-Managers were company managers with more than 10 employees or individuals with a profession requiring university training; Independent workers were individuals who worked as consultants, were independent or were company managers with fewer than 10 employees. Individuals were considered to have a prior SARS-CoV-2 infection if self-reported or if their serological test was positive for anti-SARS-CoV-2 antibodies as part of the seroprevalence studies. COVID-19 vaccination willingness was defined as the combined answer to the following two questions used in the larger vaccination questionnaire: “Did you get vaccinated against SARS-CoV-2? (Yes, No, Scheduled appointment)” and “Do you intend to get vaccinated once you will be eligible for vaccination against SARS-CoV-2? (Yes, rather yes, rather no, no, does not know, not available)”. Answers “Yes”, “Scheduled appointment” to the first question and answers “Yes” and “Rather yes” to the second question were later combined as willingness to get vaccinated. Answers “No” to the first question, and “answers No” and “Rather no” to the second question were later combined as no willingness to get vaccinated. Vaccination perception was defined as the answer to the question used in the larger vaccination questionnaire: “Do you think that vaccination is an important step to surmount the pandemic?” (Yes, rather yes, rather no, no).

Logistic regression models were used to evaluate associations between different factors and the absence of any context in which participants believed a vaccination certificate should be presented. The factors considered were age, gender, education level, household income, employment status, occupational position, past SARS-CoV-2 infection, vaccination willingness, and vaccination perception. The outcome was certificate non-acceptance, defined by the variable “There is no context in which a vaccination certificate should be presented”. Multivariable regression models were then used to calculate adjusted odds ratios (aOR) with a 95% confidence interval (95% CI). Odds ratios (aOR) were adjusted for age, gender, education level, household income, employment status, occupational position, past SARS-CoV-2 infection, vaccination willingness, and vaccination perception.

## Results

Out of 5,230 individuals, 4,056 answered the questionnaire (response rate 77.6%). Mean age (± standard deviation) was 53.3 ± 14.4 years and 56.1% were women. Overall characteristics are presented in Table 1. Non-participants (n=1,174) had a mean age of 43.7 ± 14.4 years, and 53.3% were women.

**Table 1.**
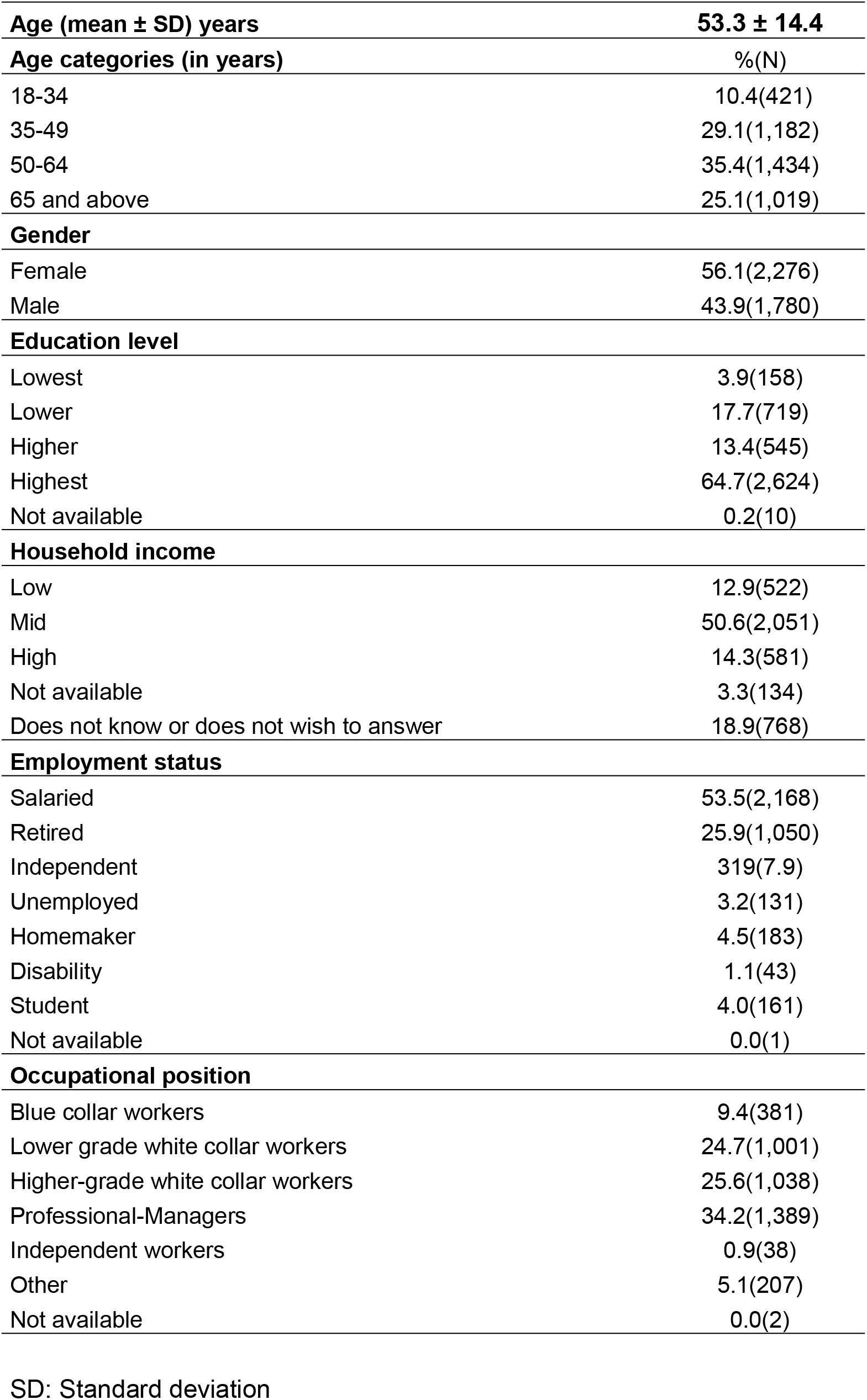
Characteristics of participants (n=4,056)

Overall, 61.0% of participants agreed or strongly agreed that a vaccination certificate is necessary in certain contexts; and 21.6% believed there was no context where vaccination certificates should be presented, defined as certificate non-acceptance (Table 2). Not willing to get vaccinated (aOR 8.07; 95% CI 6.29-10.36) and not perceiving vaccination as an important step to surmount the pandemic (aOR 3.94; 95% CI 2.71-5.73) were associated with certificate non-acceptance. Age 65 and above (aOR 0.48; 95% CI 0.31-0.73) was inversely associated with certificate non-acceptance. Past SARS-CoV-2 infection (OR 1.53; 95% CI 1.27-1.82) and female gender (OR 1.40; 95% CI 1.20-1.63) were associated with certificate non-acceptance in the univariate analyses but not when adjusted in the multivariable analyses. High household income (OR 0.68; 95% CI 0.50-0.91) and being a professional-manager (OR 0.67; 95% CI 0.51-0.89) were inversely associated with certificate non-acceptance in the univariate analyses but not when adjusted in the multivariable analyses (Table 3).

**Table 2.** Overall results with percentages out of 4,056 participants

|  | %(N) |
| --- | --- |
| **In which context(s) should a vaccination certificate be presented?** |  |
| If working in a job where others would be at risk (for ex. in nursing homes) | 60.7(2,462) |
| If working in a job where the employee is at high risk of infection (for ex. in hospitals) | 58.7(2,380) |
| If working in a job where employees have to share the same open workspace | 21.5(872) |
| To visit high risk individuals (for ex. in nursing homes or hospitals) | 47.7(1,933) |
| To cross national borders | 32.2(1,306) |
| To take a plane | 44.3(1,795) |
| To be exempt from quarantine when travelling abroad | 56.1(2,274) |
| To participate in large gatherings (for ex. concerts, matches etc.) | 36.9(1,496) |
| To have access to social venues (for ex. cinema, theater, sports club) | 35.5(1,439) |
| There is no context where a certificate should be presented | 21.6(876) |
| Other contexts | 1.4(55) |
| **What is your opinion about the following statements on a scale of 1 (Strongly disagree) to 5 (Strongly agree)?** |  |
| A vaccination certificate should be necessary in certain contexts (for ex. to travel, take care of vulnerable individuals) |  |
| Strongly disagree | 13.6(550) |
| Disagree | 8.7(353) |
| Neither agree nor disagree | 16.7(678) |
| Agree | 23.2(941) |
| Strongly agree | 37.8(1,534) |
| COVID-19 is a trivial disease that does not necessitate a vaccination certificate |  |
| Strongly disagree | 49.9(2,024) |
| Disagree | 20.5(830) |
| Neither agree nor disagree | 17.1(695) |
| Agree | 7.9(320) |
| Strongly agree | 4.6(18) |
| Individuals without a vaccination certificate could be victims of discrimination (for ex. employment opportunities, participating in activities) |  |
| Strongly disagree | 9.4(380) |
| Disagree | 9.4(383) |
| Neither agree nor disagree | 22.7(921) |
| Agree | 27.0(1,093) |
| Strongly agree | 31.5(1,279) |
| Individuals without a vaccination certificate risk losing certain rights (for ex. crossing borders) |  |
| Strongly disagree | 7.4(302) |
| Disagree | 7.5(304) |
| Neither agree nor disagree | 18.5(751) |
| Agree | 31.7(1,285) |
| Strongly agree | 34.9(1,414) |
| Personal medical data belongs to the individual and should not be the object of a vaccination certificate |  |
| Strongly disagree | 21.9(890) |
| Disagree | 15.9(645) |
| Neither agree nor disagree | 22.2(902) |
| Agree | 13.2(536) |
| Strongly agree | 26.7(1,083) |
| It is easier to accept a vaccination certificate than the public health restrictions (for ex. partial lockdown, business closures) |  |
| Strongly disagree | 8.8(356) |
| Disagree | 8.7(352) |
| Neither agree nor disagree | 20.5(833) |
| Agree | 24.7(1,000) |
| Strongly agree | 37.4(1,515) |

**Table 3.**
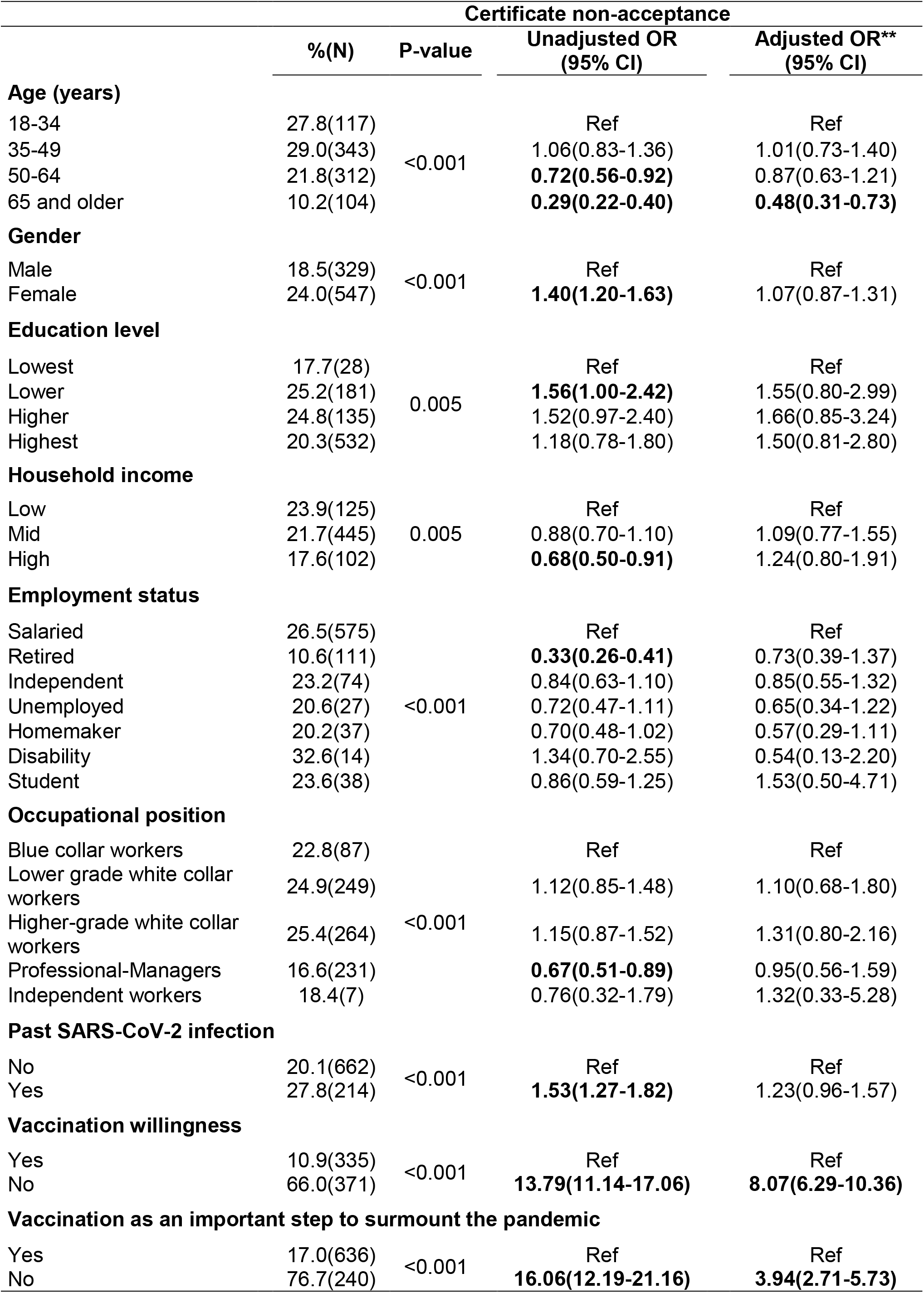
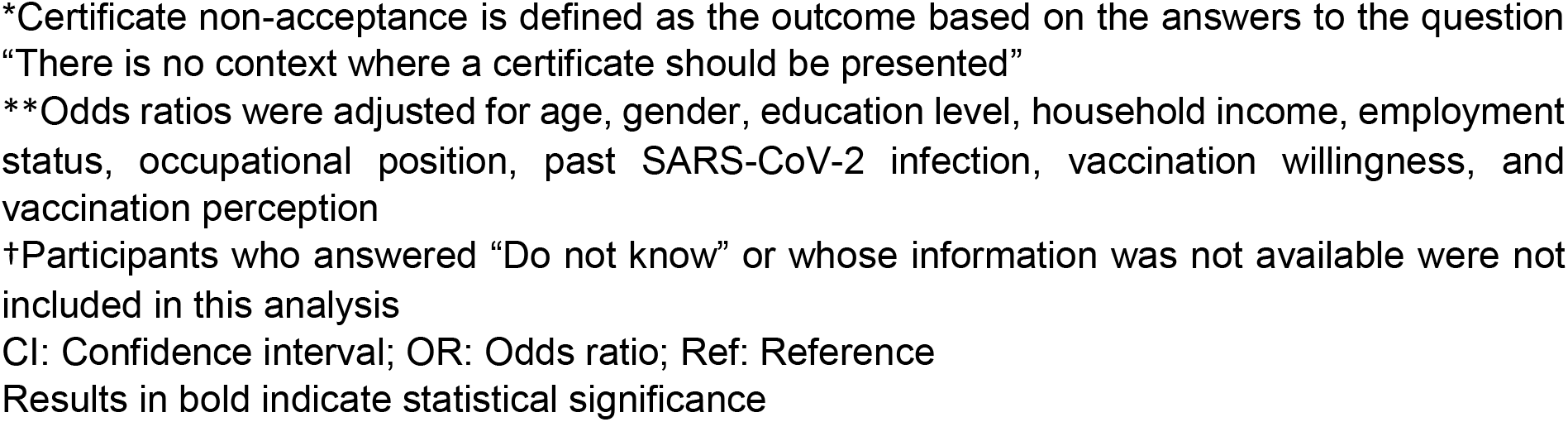
Associations between baseline characteristics, past SARS-CoV-2 infection, perception and willingness to get vaccinated, and certificate non-acceptance*†

| Certificate non-acceptance |  |  |  |  |
| --- | --- | --- | --- | --- |
|  | %(N) | P-value | Unadjusted OR<br>(95% CI) | Adjusted OR**<br>(95% CI) |
| **Age (years)** |  |  |  |  |
| 18-34 | 27.8(117) | <0.001 | Ref | Ref |
| 35-49 | 29.0(343) |  | 1.06(0.83-1.36) | 1.01(0.73-1.40) |
| 50-64 | 21.8(312) |  | **0.72(0.56-0.92)** | 0.87(0.63-1.21) |
| 65 and older | 10.2(104) |  | **0.29(0.22-0.40)** | **0.48(0.31-0.73)** |
| **Gender** |  |  |  |  |
| Male | 18.5(329) | <0.001 | Ref | Ref |
| Female | 24.0(547) |  | **1.40(1.20-1.63)** | 1.07(0.87-1.31) |
| **Education level** |  |  |  |  |
| Lowest | 17.7(28) | 0.005 | Ref | Ref |
| Lower | 25.2(181) |  | **1.56(1.00-2.42)** | 1.55(0.80-2.99) |
| Higher | 24.8(135) |  | 1.52(0.97-2.40) | 1.66(0.85-3.24) |
| Highest | 20.3(532) |  | 1.18(0.78-1.80) | 1.50(0.81-2.80) |
| **Household income** |  |  |  |  |
| Low | 23.9(125) | 0.005 | Ref | Ref |
| Mid | 21.7(445) |  | 0.88(0.70-1.10) | 1.09(0.77-1.55) |
| High | 17.6(102) |  | **0.68(0.50-0.91)** | 1.24(0.80-1.91) |
| **Employment status** |  |  |  |  |
| Salaried | 26.5(575) | <0.001 | Ref | Ref |
| Retired | 10.6(111) |  | **0.33(0.26-0.41)** | 0.73(0.39-1.37) |
| Independent | 23.2(74) |  | 0.84(0.63-1.10) | 0.85(0.55-1.32) |
| Unemployed | 20.6(27) |  | 0.72(0.47-1.11) | 0.65(0.34-1.22) |
| Homemaker | 20.2(37) |  | 0.70(0.48-1.02) | 0.57(0.29-1.11) |
| Disability | 32.6(14) |  | 1.34(0.70-2.55) | 0.54(0.13-2.20) |
| Student | 23.6(38) |  | 0.86(0.59-1.25) | 1.53(0.50-4.71) |
| **Occupational position** |  |  |  |  |
| Blue collar workers | 22.8(87) | <0.001 | Ref | Ref |
| Lower grade white collar workers | 24.9(249) |  | 1.12(0.85-1.48) | 1.10(0.68-1.80) |
| Higher-grade white collar workers | 25.4(264) |  | 1.15(0.87-1.52) | 1.31(0.80-2.16) |
| Professional-Managers | 16.6(231) |  | **0.67(0.51-0.89)** | 0.95(0.56-1.59) |
| Independent workers | 18.4(7) |  | 0.76(0.32-1.79) | 1.32(0.33-5.28) |
| **Past SARS-CoV-2 infection** |  |  |  |  |
| No | 20.1(662) | <0.001 | Ref | Ref |
| Yes | 27.8(214) |  | **1.53(1.27-1.82)** | 1.23(0.96-1.57) |
| **Vaccination willingness** |  |  |  |  |
| Yes | 10.9(335) | <0.001 | Ref | Ref |
| No | 66.0(371) |  | **13.79(11.14-17.06)** | **8.07(6.29-10.36)** |
| **Vaccination as an important step to surmount the pandemic** |  |  |  |  |
| Yes | 17.0(636) | <0.001 | Ref | Ref |
| No | 76.7(240) |  | **16.06(12.19-21.16)** | **3.94(2.71-5.73)** |

\*Certificate non-acceptance is defined as the outcome based on the answers to the question "There is no context where a certificate should be presented"
\*\*Odds ratios were adjusted for age, gender, education level, household income, employment status, occupational position, past SARS-CoV-2 infection, vaccination willingness, and vaccination perception
†Participants who answered "Do not know" or whose information was not available were not included in this analysis
CI: Confidence interval; OR: Odds ratio; Ref: Reference
Results in bold indicate statistical significance

When selecting contexts, 60.7% of participants found that a vaccination certificate should be presented in order to hold a job or position that requires contact with populations at risk of complications from COVID-19 (working in a nursing home for example), and 47.7% of participants believed a vaccination certificate should be presented when visiting individuals at risk of complications from COVID-19. Overall, 58.7% of participants found that a vaccination certificate should be presented in order to hold a job or position that required contact with infected individuals (working at a hospital for example), and 21.5% considered that a vaccination certificate should be presented if employees were sharing the same open workspace.

When considering collective versus individual benefit, 32.2% believed a vaccination certificate should be presented in order to cross international borders, 44.3% of participants believed such certificates should be presented to take a plane and 56.1% of participants believed they should be presented in order to avoid quarantine when crossing international borders. With regards to specific activities, 36.9% of participants believed vaccination certificates should be presented in order to participate in large gatherings and 35.5% in order to have access to social venues (cinema, theater, gym etc.).

Overall, 62.1% agreed or strongly agreed that it was easier to accept vaccination certificates than the public health restrictions in place at the time of the questionnaire, and 12.5% agreed or strongly agreed with the statement that COVID-19 was a trivial disease that did not require a vaccination certificate. When discussing potential risks, 39.9% of participants agreed or strongly agreed that vaccination status constituted personal medical data that should not be the object of a vaccination certificate; 58.5% of participants agreed or strongly agreed that individuals without a vaccination certificate could be at risk of discrimination (employment opportunities or participating in certain activities for example), and 66.5% agreed or strongly agreed that individuals without a vaccination certificate could lose certain rights (crossing borders for example).

Stratification by age, gender, education level, household income, employment status, occupational position, past SARS-CoV-2 infection, vaccination willingness, and vaccination perception, is presented in Appendix table 1.

### Vaccination willingness and perception

Overall, 66.0% of individuals who did not or will not get vaccinated (371 out of 562) did not believe there was any context where a vaccination certificate should be presented versus 10.9% of individuals who reported they got or intended to get vaccinated (170 out of 2,373). Additionally, participants who did not believe vaccination to be an important step in surmounting the COVID-19 pandemic rejected all contexts of vaccination certificates, with 76.7% of them seeing no context where vaccination certificates should be presented (240 out of 313) versus 17.0% of individuals who believed vaccination to be an important step in surmounting the COVID-19 pandemic (635 out of 3,743). Participants who did not believe vaccination to be an important step in surmounting the COVID-19 pandemic were also more likely to perceive a discrimination risk against individuals without a vaccination certificate, were more likely to agree that vaccination status was personal data that should not be the object of a vaccination certificate, and were less likely to agree that certificates were easier to accept than the public health restrictions in place at the time of the questionnaire.

### Past SARS-CoV-2 infection

Participants who had been infected with SARS-CoV-2 were less likely to support vaccination certificates. Overall, 27.8% of individuals who had been infected did not believe there was any context where a vaccination certificate should be presented, versus 20.1% of individuals who had not been infected. Additionally, 47.2% of previously infected individuals were likely to agree or strongly agree that vaccination status was personal medical data that should not be the object of a vaccination certificate versus 38.3% of individuals who had not been infected. Of previously infected individuals, 15.2% agreed or strongly agreed with the statement that COVID-19 was a trivial disease not necessitating a vaccination certificate versus 11.8% of non-infected individuals.

### Sociodemographic characteristics

Older individuals were more likely to agree with a vaccination certificate overall, whether in a professional, travel or social context. They were more likely to disagree with the statement that COVID-19 was a trivial disease not necessitating a vaccination certificate. Younger individuals were more likely to agree or strongly agree that individuals without vaccination certificates might lose certain rights or be at risk of discrimination.

Men were more inclined to agree with the use of vaccination certificates in all listed contexts, and when compared to the public health measures in place at the time of the questionnaire, they were also less likely to agree that individuals without vaccination certificates might face discrimination or lose certain rights. Women more strongly agreed that a vaccination status constituted personal medical data that should not be the object of a vaccination certificate.

Individuals with a high education level were more likely to agree with the statement that vaccination certificates were needed in professional settings where others would be at risk of COVID-related complications or where the employee might be at risk of infection, whereas no difference was seen in contexts of travel, access to social venues or large gatherings. Individuals with a high education level were less likely to agree with the statement that vaccination status constituted personal medical data that should not be the object of a certificate.

Individuals with a low household income agreed less overall with the use of vaccination certificates. They were more likely to agree that COVID-19 was a trivial disease not necessitating a vaccination certificate, and that vaccination status constituted personal medical data that should not be the object of a certificate. They were less likely to agree that individuals without a vaccination certificate were at risk of discrimination or may lose certain rights. They were also less likely to agree with the fact that it would be easier to accept vaccination certificates than the public health restrictions in place at the time of the questionnaire.

Managers were more inclined to agree with vaccination certificates in all contexts. More participants in the professional-managers category agreed that vaccination certificates should be presented to take a plane or to be exempt from quarantine when traveling abroad. Fewer participants in the professional-managers category agreed that vaccination status constituted personal medical data that should not be the object of a certificate.

## Discussion

Using a large population-based study, we found that 61.0% of individuals agreed or strongly agreed that a vaccination certificate is necessary in certain contexts, and 62.1% agreed or strongly agreed that it was easier to accept vaccination certificates than the public health restrictions in place at the time of the questionnaire. Certificate non-acceptance (overall 21.6%) was associated with younger age, an absence of willingness to get vaccinated and an absence of belief in vaccination as an important step in surmounting the pandemic. Overall, 27.8% of 18-34 years old reported certificate non-acceptance versus 10.2% of individuals 65 years and older. By comparison, a recent French survey (n=3,058) reported a 34.1% overall certificate non-acceptance rate (44.4% in individuals 18-34 years old versus 16.7% in individuals 65 years and older)^26^, and a UK-based public poll (n=1,715) reported a 34% overall certificate non-acceptance rate (42% in individuals 18-24 and 25-49 years old versus 20% in individuals 65 years and older)^27^.

Contexts where a majority of participants perceived a vaccination certificate should be presented included jobs where others would be at risk of COVID-related complications and jobs where employees were at risk of getting infected. This is in line with recent articles showing the need for safeguards around work-related vaccination policies that should be based on the actual risk to workers’ or customers’ health^28,29^. The majority of participants were also in favor of a vaccination certificate when presented with the option of quarantine exemption if traveling abroad, which could be a way to reinvigorate the tourism and travel sectors that have suffered greatly during the pandemic. On the other hand, participants were less in favor of a vaccination certificate in order to participate in large gatherings or to access social venues, where it might be up to private actors to decide whether vaccines are mandatory, thus potentially influencing vaccination uptake^28^. Interestingly, in a canton that borders France and where many individuals cross borders frequently, only 32.2% believed vaccination certificates should be presented to cross borders, while 44.3% believed they should be presented to take a plane. This could also be due to the perception or impression of an increased risk of infection when taking a plane versus other means of transportation. A public poll in the UK in March 2021^27^ revealed that 72% of participants believed vaccination certificates should be required to visit nursing homes, followed by gyms (56%), pubs and bars (56%), cinemas (55%), restaurants (53%), public transport (45%), and supermarkets (31%) underlining the differences in acceptability according to contexts^27^.

When addressing the potential risks around vaccination certificates, the majority of individuals perceived risks of discrimination and loss of certain rights for those without a vaccination certificate. Less than forty percent perceived vaccination status as personal medical data that should not be part of a vaccination certificate. While vaccination certificates risk infringing on civil liberties by putting pressure on individuals to share their information, it must also be recognized that lockdown measures and the pandemic itself have also represented a burden on civil liberties such as free movement or allowing people to return to work^30^. This was also evidenced in our study where a majority of participants agreed that it was easier to accept a vaccination certificate than the public health restrictions in place at the time of the questionnaire.

Women were less likely to agree with the latter statement as well as less likely to agree with vaccination certificates in all the listed contexts, indicating they were overall less in favor of vaccination certificates. Similar results were reported in a recent publication showing that female scholars were significantly less in favor of immunity certificates^18^. Gender differences were less evident when conducting multivariable analyses in our study. Individuals who had been infected were less inclined to agree with vaccination certificates. This could be due to the fact that a higher percentage of previously infected individuals agreed with the statement that COVID-19 was a trivial disease not necessitating a vaccination certificate, or to their lower perceived personal need for vaccination, having themselves acquired natural immunity.

When comparing results to our previous survey in May-June 2020^19^, individuals still believed immunity certificates were important in certain contexts but not all, while identifying potential risks of discrimination or losing certain rights. It is important to note that the risk of deliberate infection as a means to obtain a certificate should decrease with the advent of vaccination as individuals could now make the decision to get vaccinated. That being said, it is also important to take into account that universal access to vaccines remained an issue at the time of our study, especially in low and middle income countries.

Our study adds to the general body of knowledge by providing information on the importance of taking into account differences in the perception of vaccination and the disease itself when implementing vaccination certificates. While disagreements exist regarding the justification and appropriate use of vaccination certificates on ethical grounds^31,32^, the likely public uptake of such certificates is an important factor in considering their implementation. Vaccination certificates should not be a blanket solution however, and ought to be tailored to specific contexts instead.

This study has several limitations. First, the questionnaire was only available online and in French, participants were overall older, with a higher education level and a higher socioeconomic status than the overall population of Geneva, limiting the generalizability of the results. Second, the timing of the questionnaire when partial lockdown measures were still in effect in Switzerland could have influenced opinions which could change over time. Third, our study addressed vaccination certificates only, compared to the three-modality certificates deployed in Switzerland and the EU. Fourth, participants were potentially unlikely to be aware of the advantages or disadvantages of the studied intervention as is usually the case in survey research.

## Conclusion

Vaccination certificates appear to be supported by the majority of the general population in Geneva, Switzerland, especially in contexts of quarantine exemption and where work-related transmission of SARS-CoV-2 would be reduced for individuals at risk of complications or infection. Vaccination certificates are less accepted in contexts of large gatherings, access to social venues or shared workspaces. When implemented, it is important to address and communicate the role of vaccination certificates as a transition strategy in facilitating a collective phased return to pre-COVID activities by providing reassurance to individuals pursuing these activities as to their reduced risk of transmitting or acquiring an infection. Vaccination certificates should be met with a targeted implementation, adapting them to certain contexts, and modifying or cancelling them when they are no longer needed. Implementation strategies should take into consideration personal and sociodemographic variations in certificate non-acceptance, highlighting the importance of tailoring communication to younger individuals, those who may not agree with vaccination against SARS-CoV-2, and those who believe COVID-19 to be a trivial disease. Certificates are important in their role to ensure collective safety while preventing fraud. Reports of fraudulent COVID-19 related attestations have now emerged^33^, and accurate and reliable methods are important to ensure the information presented is correct, verifiable and universal. Adapting certificates to certain contexts seems to be the best available alternative to lockdown measures ensuring both personal freedom and collective welfare.

## Data Availability

Data: available by contacting Dr. Mayssam Nehme at. Data is made available upon agreement of the data provider and signature of a data transfer agreement.

## Specchio-COVID19 study group

Isabelle Arm-Vernez, Andrew S Azman, Fatim Ba, Oumar Aly Ba, Delphine Bachmann, Jean-François Balavoine, Michael Balavoine, Hélène Baysson, Lison Beigbeder, Julie Berthelot, Patrick Bleich, Gaëlle Bryand, François Chappuis, Prune Collombet, Delphine Courvoisier, Alain Cudet, Carlos de Mestral Vargas, Paola D’ippolito, Richard Dubos, Roxane Dumont, Isabella Eckerle, Nacira El Merjani, Antoine Flahault, Natalie Francioli, Marion Frangville, Idris Guessous, Séverine Harnal, Samia Hurst, Laurent Kaiser, Omar Kherad, Julien Lamour, Pierre Lescuyer, François L’Huissier, Fanny-Blanche Lombard, Andrea Jutta Loizeau, Elsa Lorthe, Chantal Martinez, Lucie Ménard, Lakshmi Menon, Ludovic Metral-Boffod, Alexandre Moulin, Mayssam Nehme, Natacha Noël, Francesco Pennacchio, Javier Perez-Saez, Giovanni Piumatti, Didier Pittet, Jane Portier, Klara M Posfay-Barbe, Géraldine Poulain, Caroline Pugin, Nick Pullen, Zo Francia Randrianandrasana, Aude Richard, Viviane Richard, Frederic Rinaldi, Jessica Rizzo, Khadija Samir, Claire Semaani, Silvia Stringhini, Stéphanie Testini, Didier Trono, Guillemette Violot, Nicolas Vuilleumier, Ania Wisniak, Sabine Yerly, MarÍa-Eugenia Zaballa

## Statement

There are no potential conflicts of interest nor significant financial contributions to this work. We confirm that the manuscript has been read and approved by all named authors and that there are no other persons who satisfied the criteria for authorship but are not listed.

We further confirm that any aspect of the work covered in this manuscript that has involved human patients has been conducted with the ethical approval of the Cantonal Research Ethics Commission of Geneva, Switzerland, and that such approvals are acknowledged within the manuscript.

## Notes

**Funding** This work was supported by the Edmond J SAFRA foundation for clinical research in internal medicine, the General Directorate of Health of the Department of Safety, Employment and Health of the canton of Geneva, the Private Foundation of the Geneva University Hospitals, the Swiss Federal Office of Public Health, the Swiss School of Public Health (Corona Immunitas Research Program) and the Fondation des Grangettes.

### Competing Interest Statement

The authors have declared no competing interest.

### Funding Statement

This work was supported by the Edmond J SAFRA foundation for clinical research in internal medicine, the General Directorate of Health of the Department of Safety, Employment and Health of the canton of Geneva, the Private Foundation of the Geneva University Hospitals, the Swiss Federal Office of Public Health, the Swiss School of Public Health (Corona Immunitas Research Program) and the Fondation des Grangettes.

### Author Declarations

Cantonal Research Ethics Commission of Geneva, Switzerland (protocol numbers CER 2020-01540 and 2020-00881)

