## Supplemental table 1 for "Perceptions of immunity and vaccination certificates among the general population in Geneva, Switzerland"

Supplement S1. Survey items on vaccination certificates

| | |
|---|---|
| **1. Select the context(s) in which a vaccination certificate should be presented:** | |
| If working in a job where others would be at risk (for ex. in nursing homes) | ☐ |
| If working in a job where the employee is at high risk of infection (for ex. in hospitals) | ☐ |
| If working in a job where employees have to share the same open workspace | ☐ |
| To visit high risk individuals (for ex. in nursing homes or hospitals) | ☐ |
| To cross national borders | ☐ |
| To take a plane | ☐ |
| To be exempt from quarantine when travelling abroad | ☐ |
| To participate in large gatherings (for ex. concerts, matches etc.) | ☐ |
| To participate in collective social activities (for ex. cinemas, theater, sports clubs) | ☐ |
| Other contexts | ☐ |
| There is no context where a certificate should be presented | ☐ |
| **2. What is your opinion about the following statements on a scale of 1 (Strongly disagree) to 5 (Strongly agree)?** | |
| A vaccination certificate should be necessary in certain contexts (for ex. to travel, take care of vulnerable individuals) | 1 “Strongly disagree”;<br>2 “Disagree”;<br>3 “Neither agree nor disagree”;<br>4” Agree”;<br>5” Strongly agree” |
| COVID-19 is a trivial disease that does not necessitate a vaccination certificate | 1 “Strongly disagree”;<br>2 “Disagree”;<br>3 “Neither agree nor disagree”;<br>4” Agree”;<br>5” Strongly agree” |
| Individuals without a vaccination certificate could be victims of discrimination (for ex. employment opportunities, participating in activities) | 1 “Strongly disagree”;<br>2 “Disagree”;<br>3 “Neither agree nor disagree”;<br>4” Agree”;<br>5” Strongly agree” |
| Individuals without a vaccination certificate risk losing certain rights (for ex. crossing borders) | 1 “Strongly disagree”;<br>2 “Disagree”;<br>3 “Neither agree nor disagree”;<br>4” Agree”;<br>5” Strongly agree” |
| Personal medical data belongs to the individual and should not be the object of a vaccination certificate | 1 “Strongly disagree”;<br>2 “Disagree”;<br>3 “Neither agree nor disagree”;<br>4” Agree”;<br>5” Strongly agree” |
| It is easier to accept a vaccination certificate than the measures imposed by the pandemic (for ex. partial lockdown, business closures) | 1 “Strongly disagree”;<br>2 “Disagree”;<br>3 “Neither agree nor disagree”;<br>4” Agree”;<br>5” Strongly agree” |
