## Appendix table 1 for "Perceptions of immunity and vaccination certificates among the general population in Geneva, Switzerland"

Appendix 1. Stratified results by age categories, gender, education level, household income, employment status, occupational position, past SARS-CoV-2 infection, vaccination willingness, and vaccination perception.

| | Age categories (years) | | | | |
|---|---|---|---|---|---|
| | **18-34** | **35-49** | **50-64** | **65 and above** | **P-value** |
| | %(N) | %(N) | %(N) | %(N) | |
| **Select the context(s) in which a vaccination certificate should be presented:** | | | | | |
| If working in a job where others would be at risk (for ex. in nursing homes) | 57.2(241) | 55.2(652) | 61.7(885) | 67.1(684) | <0.001 |
| If working in a job where the employee is at high risk of infection (for ex. in hospitals) | 51.3(216) | 51.5(609) | 60.9(874) | 66.8(681) | <0.001 |
| If working in a job where employees have to share the same open workspace | 10.5(44) | 14.5(171) | 20.5(294) | 35.6(363) | <0.001 |
| To visit high risk individuals (for ex. in nursing homes or hospitals) | 36.8(155) | 39.2(463) | 47.1(675) | 62.8(640) | <0.001 |
| To cross national borders | 21.4(90) | 23.9(282) | 30.3(435) | 49.0(499) | <0.001 |
| To take a plane | 31.4(132) | 35.4(418) | 43.0(616) | 61.7(629) | <0.001 |
| To be exempt from quarantine when travelling abroad | 45.8(193) | 47.0(555) | 55.0(788) | 72.4(738) | <0.001 |
| To participate in large gatherings (for ex. concerts, matches etc.) | 30.6(129) | 31.2(369) | 35.6(510) | 47.9(488) | <0.001 |
| To participate in collective social activities (for ex. cinemas, theater, sports clubs) | 22.6(95) | 27.2(321) | 33.4(479) | 53.4(544) | <0.001 |
| There is no context where a certificate should be presented | 27.8(117) | 29.0(343) | 21.8(312) | 10.2(104) | <0.001 |
| Other contexts | 1.0(4) | 1.4(16) | 1.5(21) | 1.4(14) | |
| What is your opinion about the following statements on a scale of 1 (Strongly disagree) to 5 (Strongly agree)? | | | | | |
| A vaccination certificate should be necessary in certain contexts (for ex. to travel, take care of vulnerable individuals) | | | | | |
| Strongly disagree | 17.8(75) | 17.5(207) | 13.5(194) | 7.3(74) | |
| Disagree | 13.8(58) | 10.7(126) | 7.8(112) | 5.6(57) | |
| Neither agree nor disagree | 23.0(97) | 20.3(240) | 15.8(227) | 11.2(114) | <0.001 |
| Agree | 22.3(94) | 24.6(291) | 24.5(351) | 20.1(205) | |
| Strongly agree | 23.0(97) | 26.9(318) | 38.4(550) | 55.8(569) | |
| COVID-19 is a trivial disease that does not necessitate a vaccination certificate | | | | | |
| Strongly disagree | 32.8(138) | 41.6(492) | 49.2(705) | 67.6(689) | |
| Disagree | 33.7(142) | 23.1(273) | 21.4(307) | 10.6(108) | |
| Neither agree nor disagree | 21.4(90) | 22.7(268) | 17.3(248) | 8.7(89) | <0.001 |
| Agree | 7.8(33) | 9.1(108) | 7.9(114) | 6.4(65) | |
| Strongly agree | 4.3(18) | 3.5(41) | 4.2(60) | 6.7(68) | |

| | | | | | |
|---|---|---|---|---|---|
| Individuals without a vaccination certificate could be victims of discrimination (for ex. employment opportunities, participating in activities) | | | | | |
| Strongly disagree | 7.1(30) | 10.5(124) | 8.8(126) | 9.8(100) | |
| Disagree | 11.6(49) | 8.0(94) | 9.4(135) | 10.3(105) | |
| Neither agree nor disagree | 16.4(69) | 19.8(234) | 22.2(318) | 29.4(300) | <0.001 |
| Agree | 29.2(123) | 26.9(318) | 28.1(403) | 24.4(249) | |
| Strongly agree | 35.6(150) | 34.9(412) | 31.5(452) | 26.0(265) | |
| Individuals without a vaccination certificate risk losing certain rights (for ex. crossing borders) | | | | | |
| Strongly disagree | 6.4(27) | 8.9(105) | 7.2(103) | 6.6(67) | |
| Disagree | 6.9(29) | 6.6(78) | 7.8(112) | 8.3(85) | |
| Neither agree nor disagree | 16.2(68) | 17.1(202) | 16.7(239) | 23.7(242) | <0.001 |
| Agree | 31.8(134) | 31.3(370) | 34.4(493) | 28.3(288) | |
| Strongly agree | 38.7(163) | 36.1(427) | 34.0(487) | 33.1(337) | |
| Personal medical data belongs to the individual and should not be the object of a vaccination certificate | | | | | |
| Strongly disagree | 11.4(48) | 14.9(176) | 21.1(302) | 35.7(364) | |
| Disagree | 15.7(66) | 14.5(171) | 17.4(249) | 15.6(159) | |
| Neither agree nor disagree | 24.0(101) | 24.0(284) | 20.8(298) | 21.5(219) | <0.001 |
| Agree | 15.0(63) | 15.1(179) | 14.2(204) | 8.8(90) | |
| Strongly agree | 34.0(143) | 31.5(372) | 26.6(381) | 18.4(187) | |
| It is easier to accept a vaccination certificate than the measures imposed by the pandemic (for ex. partial lockdown, business closures) | | | | | |
| Strongly disagree | 10.0(42) | 9.6(113) | 8.6(124) | 7.6(77) | |
| Disagree | 14.3(60) | 9.9(117) | 8.2(118) | 5.6(57) | |
| Neither agree nor disagree | 28.7(121) | 22.9(271) | 20.6(295) | 14.3(146) | <0.001 |
| Agree | 21.6(91) | 26.5(313) | 26.2(375) | 21.7(221) | |
| Strongly agree | 25.4(107) | 31.1(368) | 36.4(522) | 50.8(518) | |

| | Gender | | |
|---|---|---|---|
| | Female | Male | P-value |
| | %(N) | %(N) | |
| **Select the context(s) in which a vaccination certificate should be presented:** | | | |
| If working in a job where others would be at risk (for ex. in nursing homes) | 58.8(1,338) | 63.1(1,124) | 0.005 |
| If working in a job where the employee is at high risk of infection (for ex. in hospitals) | 56.9(1,295) | 61.0(1,085) | 0.010 |
| If working in a job where employees have to share the same open workspace | 17.5(399) | 26.6(473) | <0.001 |
| To visit high risk individuals (for ex. in nursing homes or hospitals) | 44.4(1,010) | 51.9(923) | <0.001 |
| To cross national borders | 28.9(658) | 36.4(648) | <0.001 |
| To take a plane | 39.8(906) | 49.9(889) | <0.001 |
| To be exempt from quarantine when travelling abroad | 51.8(1,178) | 61.6(1,096) | <0.001 |
| To participate in large gatherings (for ex. concerts, matches etc.) | 33.7(767) | 41.0(729) | <0.001 |
| To participate in collective social activities (for ex. cinemas, theater, sports clubs) | 31.2(711) | 40.9(728) | <0.001 |
| There is no context where a certificate should be presented | 24.0(547) | 18.5(329) | <0.001 |
| Other contexts | 1.2(28) | 1.5(27) | |
| What is your opinion about the following statements on a scale of 1 (Strongly disagree) to 5 (Strongly agree)? | | | |
| A vaccination certificate should be necessary in certain contexts (for ex. to travel, take care of vulnerable individuals) | | | |
| Strongly disagree | 15.3(348) | 11.3(202) | |
| Disagree | 9.1(207) | 8.2(146) | |
| Neither agree nor disagree | 18.1(413) | 14.9(265) | <0.001 |
| Agree | 22.9(522) | 23.5(419) | |
| Strongly agree | 34.5(786) | 42.0(748) | |
| COVID-19 is a trivial disease that does not necessitate a vaccination certificate | | | |
| Strongly disagree | 47.9(1,091) | 52.4(933) | |
| Disagree | 20.7(472) | 20.1(358) | |
| Neither agree nor disagree | 18.3(416) | 15.7(279) | 0.041 |
| Agree | 8.4(192) | 7.2(128) | |
| Strongly agree | 4.6(105) | 4.6(82) | |
| Individuals without a vaccination certificate could be victims of discrimination (for ex. employment opportunities, participating in activities) | | | |
| Strongly disagree | 9.4(215) | 9.3(165) | 0.008 |

| | | | |
|---|---|---|---|
| Disagree | 8.7(199) | 10.3(184) | |
| Neither agree nor disagree | 23.2(527) | 22.1(394) | |
| Agree | 25.3(576) | 29.0(517) | |
| Strongly agree | 33.3(759) | 29.2(520) | |
| Individuals without a vaccination certificate risk losing certain rights (for ex. crossing borders) | | | |
| Strongly disagree | 8.0(181) | 6.8(121) | |
| Disagree | 7.2(165) | 7.8(139) | |
| Neither agree nor disagree | 18.8(429) | 18.1(322) | 0.057 |
| Agree | 30.0(682) | 33.9(603) | |
| Strongly agree | 36.0(819) | 33.4(595) | |
| Personal medical data belongs to the individual and should not be the object of a vaccination certificate | | | |
| Strongly disagree | 18.8(427) | 26.0(463) | |
| Disagree | 13.0(295) | 19.7(350) | |
| Neither agree nor disagree | 23.7(539) | 20.4(363) | <0.001 |
| Agree | 14.1(321) | 12.1(215) | |
| Strongly agree | 30.5(694) | 21.9(389) | |
| It is easier to accept a vaccination certificate than the measures imposed by the pandemic (for ex. partial lockdown, business closures) | | | |
| Strongly disagree | 9.6(218) | 7.8(138) | |
| Disagree | 10.0(228) | 7.0(124) | |
| Neither agree nor disagree | 21.7(495) | 19.0(338) | <0.001 |
| Agree | 23.5(535) | 26.1(465) | |
| Strongly agree | 35.1(800) | 40.2(715) | |

| | Education | | | | | |
|---|---|---|---|---|---|---|
| | Lowest | Lower | Higher | Highest | Not available | P-value |
| | %(N) | %(N) | %(N) | %(N) | %(N) | |
| **Select the context(s) in which a vaccination certificate should be presented:** | | | | | | |
| If working in a job where others would be at risk (for ex. in nursing homes) | 58.9(93) | 53.3(383) | 56.3(307) | 63.7(1,672) | 70.0(7) | <0.001 |
| If working in a job where the employee is at high risk of infection (for ex. in hospitals) | 55.7(88) | 52.3(376) | 53.0(289) | 61.7(1,620) | 70.0(7) | <0.001 |
| If working in a job where employees have to share the same open workspace | 26.6(42) | 19.6(141) | 19.3(105) | 22.1(581) | 30.0(3) | 0.157 |
| To visit high risk individuals (for ex. in nursing homes or hospitals) | 50.0(79) | 42.7(307) | 45.5(248) | 49.4(1,295) | 40.0(4) | 0.019 |
| To cross national borders | 39.2(62) | 34.4(247) | 30.8(168) | 31.5(826) | 30.0(3) | 0.184 |
| To take a plane | 50.0(79) | 44.1(317) | 40.7(222) | 44.6(1,171) | 60.0(6) | 0.197 |
| To be exempt from quarantine when travelling abroad | 50.0(79) | 52.3(376) | 53.2(290) | 58.1(1,524) | 50.0(5) | 0.012 |
| To participate in large gatherings (for ex. concerts, matches etc.) | 34.8(55) | 33.8(243) | 34.1(186) | 38.4(1,008) | 40.0(4) | 0.101 |
| To access social venues (for ex. cinema, theater, sports club) | 36.1(57) | 35.9(258) | 31.2(170) | 36.3(952) | 20.0(2) | 0.181 |
| There is no context where a certificate should be presented | 17.7(28) | 25.2(181) | 24.8(135) | 20.3(532) | 0.0(0) | 0.003 |
| Other contexts | 1.9(3) | 1.4(10) | 1.7(9) | 1.2(32) | 10.0(1) | |
| What is your opinion about the following statements on a scale of 1 (Strongly disagree) to 5 (Strongly agree)? | | | | | | |
| A vaccination certificate should be necessary in certain contexts (for ex. to travel, take care of vulnerable individuals) | | | | | | |
| Strongly disagree | 15.2(24) | 14.9(107) | 16.3(89) | 12.6(330) | 0.0(0) | |
| Disagree | 7.6(12) | 9.3(67) | 9.0(49) | 8.6(225) | 0.0(0) | |
| Neither agree nor disagree | 16.5(26) | 17.8(128) | 20.2(110) | 15.7(411) | 30.0(3) | 0.014 |
| Agree | 20.3(32) | 19.1(137) | 22.0(120) | 24.8(650) | 20.0(2) | |
| Strongly agree | 40.5(64) | 38.9(280) | 32.5(177) | 38.4(1,008) | 50.0(5) | |
| COVID-19 is a trivial disease that does not necessitate a vaccination certificate | | | | | | |
| Strongly disagree | 43.0(68) | 46.3(333) | 46.8(255) | 51.9(1,362) | 60.0(6) | |
| Disagree | 17.1(27) | 17.7(127) | 20.4(111) | 21.5(563) | 20.0(2) | |
| Neither agree nor disagree | 18.4(29) | 19.2(138) | 20.0(109) | 15.9(418) | 10.0(1) | <0.001 |
| Agree | 10.8(17) | 10.0(72) | 7.7(42) | 7.2(189) | 0.0(0) | |
| Strongly agree | 10.8(17) | 6.8(49) | 5.1(28) | 3.5(92) | 10.0(1) | |
| Individuals without a vaccination certificate could be victims of discrimination (for ex. employment opportunities, participating in activities) | | | | | | |
| Strongly disagree | 18.4(29) | 10.2(73) | 9.4(51) | 8.5(224) | 30.0(3) | 0.001 |

| | | | | | | |
|---|---|---|---|---|---|---|
| Disagree | 12.0(19) | 10.2(73) | 9.0(49) | 9.2(242) | 0.0(0) | |
| Neither agree nor disagree | 21.5(34) | 22.1(159) | 21.7(118) | 23.1(607) | 30.0(3) | |
| Agree | 17.7(28) | 23.2(167) | 27.9(152) | 28.4(744) | 20.0(2) | |
| Strongly agree | 30.4(48) | 34.4(247) | 32.1(175) | 30.8(807) | 20.0(2) | |
| Individuals without a vaccination certificate risk losing certain rights (for ex. crossing borders) | | | | | | |
| Strongly disagree | 12.0(19) | 8.2(59) | 7.3(40) | 6.9(182) | 20.0(2) | |
| Disagree | 12.0(19) | 7.6(55) | 8.1(44) | 7.1(186) | 0.0(0) | |
| Neither agree nor disagree | 20.3(32) | 19.1(137) | 18.9(103) | 18.1(476) | 30.0(3) | 0.005 |
| Agree | 19.0(30) | 27.3(196) | 31.7(173) | 33.7(884) | 20.0(2) | |
| Strongly agree | 36.7(58) | 37.8(272) | 33.9(185) | 34.1(896) | 30.0(3) | |
| Personal medical data belongs to the individual and should not be the object of a vaccination certificate | | | | | | |
| Strongly disagree | 15.2(24) | 20.4(147) | 19.6(107) | 23.2(608) | 40.0(4) | |
| Disagree | 12.7(20) | 10.4(75) | 13.6(74) | 18.1(476) | 0.0(0) | |
| Neither agree nor disagree | 27.8(44) | 23.6(170) | 21.8(119) | 21.6(566) | 30.0(3) | <0.001 |
| Agree | 12.7(20) | 13.4(96) | 15.2(83) | 12.8(336) | 10.0(1) | |
| Strongly agree | 31.6(50) | 32.1(231) | 29.7(162) | 24.3(638) | 20.0(2) | |
| It is easier to accept a vaccination certificate than the measures imposed by the pandemic (for ex. partial lockdown, business closures) | | | | | | |
| Strongly disagree | 12.7(20) | 10.7(77) | 9.9(54) | 7.8(205) | 0.0(0) | |
| Disagree | 8.9(14) | 7.6(55) | 10.6(58) | 8.5(224) | 10.0(1) | |
| Neither agree nor disagree | 21.5(34) | 23.6(170) | 24.4(133) | 18.8(493) | 30.0(3) | <0.001 |
| Agree | 15.8(25) | 21.0(151) | 23.5(128) | 26.5(695) | 10.0(1) | |
| Strongly agree | 41.1(65) | 37.0(266) | 31.6(172) | 38.4(1,007) | 50.0(5) | |

| | Household income | | | | | |
|---|---|---|---|---|---|---|
| | Low | Mid | High | Does not know or does not wish to answer | Not available | P-value |
| | %(N) | %(N) | %(N) | %(N) | %(N) | |
| **Select the context(s) in which a vaccination certificate should be presented:** | | | | | | |
| If working in a job where others would be at risk (for ex. in nursing homes) | 59.2(309) | 60.5(1,241) | 66.3(385) | 59.2(455) | 53.7(72) | 0.022 |
| If working in a job where the employee is at high risk of infection (for ex. in hospitals) | 54.2(283) | 59.3(1,217) | 63.0(366) | 58.1(446) | 50.7(68) | 0.013 |
| If working in a job where employees have to share the same open workspace | 24.9(130) | 21.5(440) | 21.3(124) | 20.7(159) | 14.2(19) | 0.087 |
| To visit high risk individuals (for ex. in nursing homes or hospitals) | 47.3(247) | 47.1(967) | 52.7(306) | 47.3(363) | 37.3(50) | 0.018 |
| To cross national borders | 35.6(186) | 31.1(638) | 34.9(203) | 31.8(244) | 26.1(35) | 0.083 |
| To take a plane | 42.5(222) | 43.5(893) | 49.2(286) | 45.7(351) | 32.1(43) | 0.004 |
| To be exempt from quarantine when travelling abroad | 51.5(269) | 56.1(1,150) | 64.0(372) | 55.7(428) | 41.0(55) | <0.001 |
| To participate in large gatherings (for ex. concerts, matches etc.) | 35.1(183) | 36.1(740) | 44.8(260) | 35.0(269) | 32.8(44) | 0.001 |
| To participate in collective social activities (for ex. cinemas, theater, sports clubs) | 35.4(185) | 34.9(715) | 42.2(245) | 34.2(263) | 23.1(31) | <0.001 |
| There is no context where a certificate should be presented | 23.9(125) | 21.7(445) | 17.6(102) | 21.1(162) | 31.3(42) | 0.005 |
| Other contexts | 0.8(4) | 1.5(30) | 1.2(7) | 1.7(13) | 0.7(1) | 0.616 |
| What is your opinion about the following statements on a scale of 1 (Strongly disagree) to 5 (Strongly agree)? | | | | | | |
| A vaccination certificate should be necessary in certain contexts (for ex. to travel, take care of vulnerable individuals) | | | | | | |
| Strongly disagree | 15.9(83) | 13.0(267) | 11.4(66) | 14.1(108) | 19.4(26) | <0.001 |
| Disagree | 9.8(51) | 8.6(177) | 6.9(40) | 9.8(75) | 7.5(10) | |
| Neither agree nor disagree | 17.4(91) | 16.0(329) | 12.7(74) | 20.3(156) | 20.9(28) | |
| Agree | 21.6(113) | 24.6(505) | 26.5(154) | 18.8(144) | 18.7(25) | |
| Strongly agree | 35.2(184) | 37.7(773) | 42.5(247) | 37.1(285) | 33.6(45) | |
| COVID-19 is a trivial disease that does not necessitate a vaccination certificate | | | | | | |
| Strongly disagree | 46.2(241) | 51.3(1,052) | 54.6(317) | 47.0(361) | 39.6(53) | <0.001 |
| Disagree | 18.0(94) | 20.0(411) | 21.3(124) | 21.1(162) | 29.1(39) | |

| | | | | | | |
|---|---|---|---|---|---|---|
| Neither agree nor disagree | 18.6(97) | 16.8(344) | 14.1(82) | 19.5(150) | 16.4(22) | |
| Agree | 10.2(53) | 8.0(164) | 7.1(41) | 6.8(52) | 7.5(10) | |
| Strongly agree | 7.1(37) | 3.9(80) | 2.9(17) | 5.6(43) | 7.5(10) | |
| Individuals without a vaccination certificate could be victims of discrimination (for ex. employment opportunities, participating in activities) | | | | | | |
| Strongly disagree | 10.0(52) | 8.7(179) | 9.6(56) | 11.1(85) | 6.0(8) | |
| Disagree | 10.2(53) | 8.7(179) | 10.5(61) | 10.5(81) | 6.7(9) | |
| Neither agree nor disagree | 23.4(122) | 24.2(497) | 20.1(117) | 21.4(164) | 15.7(21) | <0.001 |
| Agree | 20.5(107) | 26.9(551) | 33.0(192) | 25.0(192) | 38.1(51) | |
| Strongly agree | 36.0(188) | 31.4(645) | 26.7(155) | 32.0(246) | 33.6(45) | |
| Individuals without a vaccination certificate risk losing certain rights (for ex. crossing borders) | | | | | | |
| Strongly disagree | 10.2(53) | 6.5(133) | 7.2(42) | 8.6(66) | 6.0(8) | |
| Disagree | 6.7(35) | 7.5(154) | 7.2(42) | 8.2(63) | 7.5(10) | |
| Neither agree nor disagree | 18.2(95) | 19.2(393) | 16.7(97) | 20.1(154) | 9.0(12) | 0.001 |
| Agree | 27.4(143) | 32.3(662) | 37.7(219) | 27.3(210) | 38.1(51) | |
| Strongly agree | 37.5(196) | 34.6(709) | 31.2(181) | 35.8(275) | 39.6(53) | |
| Personal medical data belongs to the individual and should not be the object of a vaccination certificate | | | | | | |
| Strongly disagree | 15.5(81) | 22.4(460) | 28.6(166) | 19.8(152) | 23.1(31) | |
| Disagree | 11.7(61) | 17.2(352) | 19.6(114) | 13.3(102) | 11.9(16) | |
| Neither agree nor disagree | 25.9(135) | 21.6(444) | 20.3(118) | 22.9(176) | 21.6(29) | <0.001 |
| Agree | 12.5(65) | 13.0(266) | 13.1(76) | 14.5(111) | 13.4(18) | |
| Strongly agree | 34.5(180) | 25.8(529) | 18.4(107) | 29.6(227) | 29.9(40) | |
| It is easier to accept a vaccination certificate than the measures imposed by the pandemic (for ex. partial lockdown, business closures) | | | | | | |
| Strongly disagree | 12.3(64) | 8.2(168) | 6.9(40) | 8.5(65) | 14.2(19) | |
| Disagree | 7.7(40) | 8.3(170) | 7.1(41) | 10.8(83) | 13.4(18) | |
| Neither agree nor disagree | 23.0(120) | 20.4(419) | 13.6(79) | 23.4(180) | 26.1(35) | <0.001 |
| Agree | 23.2(121) | 25.5(522) | 26.9(156) | 21.9(168) | 24.6(33) | |
| Strongly agree | 33.9(177) | 37.6(772) | 45.6(265) | 35.4(272) | 21.6(29) | |

| | Employment status | | | | | | | | |
|---|---|---|---|---|---|---|---|---|---|
| | Salaried | Retired | Independent | Unemployed | Homemaker | Disability | Student | Not available | P-value |
| | %(N) | %(N) | %(N) | %(N) | %(N) | %(N) | %(N) | %(N) | |
| **Select the context(s) in which a vaccination certificate should be presented:** | | | | | | | | | |
| If working in a job where others would be at risk (for ex. in nursing homes) | 58.2(1,261) | 66.1(694) | 60.2(192) | 58.8(77) | 62.3(114) | 46.5(20) | 64.0(103) | 100.0(1) | <0.001 |
| If working in a job where the employee is at high risk of infection (for ex. in hospitals) | 55.0(1,193) | 66.4(697) | 58.3(186) | 58.8(77) | 60.1(110) | 46.5(20) | 59.6(96) | 100.0(1) | <0.001 |
| If working in a job where employees have to share the same open workspace | 16.1(350) | 34.2(359) | 20.4(65) | 22.9(30) | 21.3(39) | 18.6(8) | 12.4(20) | 100.0(1) | <0.001 |
| To visit high risk individuals (for ex. in nursing homes or hospitals) | 41.8(906) | 60.8(638) | 46.7(149) | 45.0(59) | 53.0(97) | 39.5(17) | 41.0(66) | 100.0(1) | <0.001 |
| To cross national borders | 25.8(559) | 48.4(508) | 28.8(92) | 27.5(36) | 32.8(60) | 27.9(12) | 23.6(38) | 100.0(1) | <0.001 |
| To take a plane | 37.0(802) | 61.5(646) | 43.3(138) | 43.5(57) | 47.0(86) | 30.2(13) | 32.3(52) | 100.0(1) | <0.001 |
| To be exempt from quarantine when travelling abroad | 50.3(1,091) | 72.0(756) | 54.9(175) | 45.8(60) | 53.6(98) | 37.2(16) | 47.8(77) | 100.0(1) | <0.001 |
| To participate in large gatherings (for ex. concerts, matches etc.) | 31.9(692) | 47.7(501) | 36.4(116) | 35.1(46) | 38.3(70) | 30.2(13) | 35.4(57) | 100.0(1) | <0.001 |
| To participate in collective social activities (for ex. cinemas, theater, sports clubs) | 28.9(627) | 52.1(547) | 33.2(106) | 31.3(41) | 36.1(66) | 32.6(14) | 23.0(37) | 100.0(1) | <0.001 |
| There is no context where a certificate should be presented | 26.5(575) | 10.6(111) | 23.2(74) | 20.6(27) | 20.2(37) | 32.6(14) | 23.6(38) | 0.0(0) | <0.001 |
| Other contexts | 1.4(31) | 1.5(16) | 0.9(3) | 0.8(1) | 0.5(1) | 0.0(0) | 1.9(3) | 0.0(0) | |
| What is your opinion about the following statements on a scale of 1 (Strongly disagree) to 5 (Strongly agree)? | | | | | | | | | |
| A vaccination certificate should be necessary in certain contexts (for ex. to travel, take care of vulnerable individuals) | | | | | | | | | |
| Strongly disagree | 15.7(341) | 7.0(73) | 15.7(50) | 16.0(21) | 17.5(32) | 20.9(9) | 14.9(24) | 0.0(0) | <0.001 |
| Disagree | 10.2(221) | 6.0(63) | 9.7(31) | 6.9(9) | 3.8(7) | 11.6(5) | 10.6(17) | 0.0(0) | |
| Neither agree nor disagree | 18.0(390) | 10.9(114) | 15.0(48) | 24.4(32) | 19.1(35) | 23.3(10) | 30.4(49) | 0.0(0) | |
| Agree | 24.7(536) | 20.6(216) | 24.5(78) | 20.6(27) | 20.8(38) | 20.9(9) | 22.4(36) | 100.0(1) | |
| Strongly agree | 31.4(680) | 55.6(584) | 35.1(112) | 32.1(42) | 38.8(71) | 23.3(10) | 21.7(35) | 0.0(0) | |
| COVID-19 is a trivial disease that does not necessitate a vaccination certificate | | | | | | | | | |
| Strongly disagree | 44.2(958) | 66.5(698) | 48.0(153) | 43.5(57) | 49.7(91) | 39.5(17) | 30.4(49) | 100.0(1) | <0.001 |
| Disagree | 23.0(498) | 11.0(116) | 23.5(75) | 22.1(29) | 20.2(37) | 18.6(8) | 41.6(67) | 0.0(0) | |

| | | | | | | | | | |
|---|---|---|---|---|---|---|---|---|---|
| Neither agree nor disagree | 20.9(454) | 9.0(95) | 16.3(52) | 22.9(30) | 13.7(25) | 20.9(9) | 18.6(30) | 0.0(0) | |
| Agree | 8.4(182) | 6.8(71) | 8.5(27) | 6.9(9) | 9.8(18) | 9.3(4) | 5.6(9) | 0.0(0) | |
| Strongly agree | 3.5(76) | 6.7(70) | 3.8(12) | 4.6(6) | 6.6(12) | 11.6(5) | 3.7(6) | 0.0(0) | |
| Individuals without a vaccination certificate could be victims of discrimination (for ex. employment opportunities, participating in activities) | | | | | | | | | |
| Strongly disagree | 8.9(192) | 9.3(98) | 10.0(32) | 9.9(13) | 14.8(27) | 9.3(4) | 8.7(14) | 0.0(0) | |
| Disagree | 8.8(191) | 10.1(106) | 12.5(40) | 6.9(9) | 8.7(16) | 4.7(2) | 11.2(18) | 100.0(1) | |
| Neither agree nor disagree | 20.4(442) | 29.2(307) | 20.7(66) | 23.7(31) | 21.9(40) | 34.9(15) | 12.4(20) | 0.0(0) | <0.001 |
| Agree | 28.4(615) | 24.1(253) | 28.8(92) | 26.7(35) | 21.3(39) | 18.6(8) | 31.7(51) | 0.0(0) | |
| Strongly agree | 33.6(728) | 27.2(286) | 27.9(89) | 32.8(43) | 33.3(61) | 32.6(14) | 36.0(58) | 0.0(0) | |
| Individuals without a vaccination certificate risk losing certain rights (for ex. crossing borders) | | | | | | | | | |
| Strongly disagree | 7.4(160) | 6.0(63) | 8.5(27) | 7.6(10) | 14.2(26) | 4.7(2) | 8.1(13) | 100.0(1) | |
| Disagree | 6.9(150) | 8.1(85) | 9.4(30) | 6.9(9) | 8.2(15) | 11.6(5) | 6.2(10) | 0.0(0) | |
| Neither agree nor disagree | 16.3(353) | 23.2(244) | 20.7(66) | 18.3(24) | 16.4(30) | 23.3(10) | 14.9(24) | 0.0(0) | <0.001 |
| Agree | 33.3(723) | 29.4(309) | 31.7(101) | 31.3(41) | 26.2(48) | 20.9(9) | 33.5(54) | 0.0(0) | |
| Strongly agree | 36.1(782) | 33.2(349) | 29.8(95) | 35.9(47) | 35.0(64) | 39.5(17) | 37.3(60) | 0.0(0) | |
| Personal medical data belongs to the individual and should not be the object of a vaccination certificate | | | | | | | | | |
| Strongly disagree | 17.4(378) | 34.5(362) | 21.6(69) | 10.7(14) | 21.9(40) | 11.6(5) | 13.0(21) | 100.0(1) | |
| Disagree | 16.4(356) | 16.0(168) | 14.7(47) | 13.7(18) | 13.7(25) | 14.0(6) | 15.5(25) | 0.0(0) | |
| Neither agree nor disagree | 21.9(475) | 21.2(223) | 23.2(74) | 25.2(33) | 24.0(44) | 18.6(8) | 28.0(45) | 0.0(0) | <0.001 |
| Agree | 14.3(310) | 10.1(106) | 11.9(38) | 17.6(23) | 14.8(27) | 16.3(7) | 15.5(25) | 0.0(0) | |
| Strongly agree | 29.9(649) | 18.2(191) | 28.5(91) | 32.8(43) | 25.7(47) | 39.5(17) | 28.0(45) | 0.0(0) | |
| It is easier to accept a vaccination certificate than the measures imposed by the pandemic (for ex. partial lockdown, business closures) | | | | | | | | | |
| Strongly disagree | 8.5(185) | 7.6(80) | 9.4(30) | 12.2(16) | 12.0(22) | 16.3(7) | 9.9(16) | 0.0(0) | |
| Disagree | 9.9(215) | 5.4(57) | 7.2(23) | 11.5(15) | 8.2(15) | 9.3(4) | 14.3(23) | 0.0(0) | |
| Neither agree nor disagree | 22.4(486) | 13.8(145) | 19.4(62) | 27.5(36) | 22.4(41) | 37.2(16) | 29.2(47) | 0.0(0) | <0.001 |
| Agree | 26.3(571) | 22.3(234) | 29.5(94) | 19.8(26) | 18.0(33) | 16.3(7) | 21.7(35) | 0.0(0) | |
| Strongly agree | 32.8(711) | 50.9(534) | 34.5(110) | 29.0(38) | 39.3(72) | 20.9(9) | 24.8(40) | 100.0(1) | |

| | Occupational position | | | | | | | |
|---|---|---|---|---|---|---|---|---|
| | **Blue collar workers** | **Lower grade white collar workers** | **Higher grade white collar workers** | **Professional-Managers** | **Independent workers** | **Other** | **Not available** | **P-value** |
| | %(N) | %(N) | %(N) | %(N) | %(N) | %(N) | %(N) | |
| **Select the context(s) in which a vaccination certificate should be presented:** | | | | | | | | |
| If working in a job where others would be at risk (for ex. in nursing homes) | 58.8(224) | 56.2(563) | 56.4(585) | 67.5(937) | 55.3(21) | 63.8(132) | 0.0(0) | <0.001 |
| If working in a job where the employee is at high risk of infection (for ex. in hospitals) | 58.0(221) | 53.9(540) | 54.9(570) | 65.4(908) | 50.0(19) | 58.9(122) | 0.0(0) | <0.001 |
| If working in a job where employees have to share the same open workspace | 25.7(98) | 19.7(197) | 17.9(186) | 24.9(346) | 26.3(10) | 16.9(35) | 0.0(0) | <0.001 |
| To visit high risk individuals (for ex. in nursing homes or hospitals) | 47.5(181) | 45.3(453) | 42.0(436) | 53.6(744) | 57.9(22) | 46.9(97) | 0.0(0) | <0.001 |
| To cross national borders | 38.8(148) | 30.8(308) | 27.3(283) | 35.2(489) | 31.6(12) | 31.9(66) | 0.0(0) | <0.001 |
| To take a plane | 43.8(167) | 41.4(414) | 39.4(409) | 50.1(696) | 47.4(18) | 44.0(91) | 0.0(0) | <0.001 |
| To be exempt from quarantine when travelling abroad | 50.1(191) | 51.6(517) | 51.5(535) | 64.2(892) | 73.7(28) | 53.6(111) | 0.0(0) | <0.001 |
| To participate in large gatherings (for ex. concerts, matches etc.) | 33.6(128) | 32.5(325) | 32.5(337) | 43.7(607) | 47.4(18) | 39.1(81) | 0.0(0) | <0.001 |
| To participate in collective social activities (for ex. cinemas, theater, sports clubs) | 35.2(134) | 33.9(339) | 30.6(318) | 40.5(562) | 47.4(18) | 32.9(68) | 0.0(0) | <0.001 |
| There is no context where a certificate should be presented | 22.8(87) | 24.9(249) | 25.4(264) | 16.6(231) | 18.4(7) | 17.4(36) | 100.0(2) | <0.001 |
| Other contexts | 1.8(7) | 1.1(11) | 1.3(13) | 1.4(20) | 0.0(0) | 1.9(4) | 0.0(0) | |
| What is your opinion about the following statements on a scale of 1 (Strongly disagree) to 5 (Strongly agree)? | | | | | | | | |
| A vaccination certificate should be necessary in certain contexts (for ex. to travel, take care of vulnerable individuals) | | | | | | | | |
| Strongly disagree | 18.1(69) | 15.8(158) | 14.8(154) | 10.3(143) | 5.3(2) | 11.6(24) | 0.0(0) | |
| Disagree | 5.2(20) | 10.2(102) | 10.1(105) | 7.4(103) | 10.5(4) | 8.2(17) | 100.0(2) | |
| Neither agree nor disagree | 19.9(76) | 15.2(152) | 19.1(198) | 14.0(195) | 13.2(5) | 25.1(52) | 0.0(0) | <0.001 |
| Agree | 19.4(74) | 22.8(228) | 22.2(230) | 25.3(351) | 28.9(11) | 22.7(47) | 0.0(0) | |
| Strongly agree | 37.3(142) | 36.1(361) | 33.8(351) | 43.0(597) | 42.1(16) | 32.4(67) | 0.0(0) | |
| COVID-19 is a trivial disease that does not necessitate a vaccination certificate | | | | | | | | |
| Strongly disagree | 44.6(170) | 47.1(471) | 44.9(466) | 58.0(806) | 52.6(20) | 44.0(91) | 0.0(0) | <0.001 |

| | | | | | | | | |
|---|---|---|---|---|---|---|---|---|
| Disagree | 13.9(53) | 19.2(192) | 22.7(236) | 19.7(273) | 21.1(8) | 31.9(66) | 100.0(2) | |
| Neither agree nor disagree | 21.3(81) | 20.3(203) | 17.1(178) | 14.3(198) | 13.2(5) | 14.5(30) | 0.0(0) | |
| Agree | 11.0(42) | 8.3(83) | 9.3(97) | 5.8(80) | 13.2(5) | 6.3(13) | 0.0(0) | |
| Strongly agree | 9.2(35) | 5.2(52) | 5.9(61) | 2.3(32) | 0.0(0) | 3.4(7) | 0.0(0) | |
| Individuals without a vaccination certificate could be victims of discrimination (for ex. employment opportunities, participating in activities) | | | | | | | | |
| Strongly disagree | 15.7(60) | 10.1(101) | 7.1(74) | 8.9(123) | 7.9(3) | 9.2(19) | 0.0(0) | |
| Disagree | 7.9(30) | 10.0(100) | 8.0(83) | 10.4(145) | 10.5(4) | 10.1(21) | 0.0(0) | |
| Neither agree nor disagree | 22.8(87) | 21.6(216) | 24.0(249) | 22.8(317) | 23.7(9) | 20.3(42) | 50.0(1) | <0.001 |
| Agree | 19.9(76) | 24.7(247) | 25.8(268) | 30.6(425) | 39.5(15) | 29.5(61) | 50.0(1) | |
| Strongly agree | 33.6(128) | 33.7(337) | 35.1(364) | 27.3(379) | 18.4(7) | 30.9(64) | 0.0(0) | |
| Individuals without a vaccination certificate risk losing certain rights (for ex. crossing borders) | | | | | | | | |
| Strongly disagree | 12.9(49) | 8.2(82) | 6.3(65) | 6.5(90) | 5.3(2) | 6.8(14) | 0.0(0) | |
| Disagree | 7.9(30) | 7.8(78) | 6.6(68) | 7.8(109) | 7.9(3) | 7.7(16) | 0.0(0) | |
| Neither agree nor disagree | 20.5(78) | 17.7(177) | 19.2(199) | 18.3(254) | 26.3(10) | 15.9(33) | 0.0(0) | <0.001 |
| Agree | 25.2(96) | 29.2(292) | 31.0(322) | 35.1(487) | 39.5(15) | 34.3(71) | 100.0(2) | |
| Strongly agree | 33.6(128) | 37.2(372) | 37.0(384) | 32.3(449) | 21.1(8) | 35.3(73) | 0.0(0) | |
| Personal medical data belongs to the individual and should not be the object of a vaccination certificate | | | | | | | | |
| Strongly disagree | 16.8(64) | 18.3(183) | 17.3(180) | 29.8(414) | 26.3(10) | 18.8(39) | 0.0(0) | |
| Disagree | 11.3(43) | 13.0(130) | 14.8(154) | 19.9(277) | 18.4(7) | 16.4(34) | 0.0(0) | |
| Neither agree nor disagree | 23.9(91) | 24.1(241) | 23.3(242) | 18.8(261) | 23.7(9) | 27.5(57) | 50.0(1) | <0.001 |
| Agree | 12.3(47) | 14.0(140) | 14.5(151) | 12.0(167) | 7.9(3) | 13.5(28) | 0.0(0) | |
| Strongly agree | 35.7(136) | 30.7(307) | 30.0(311) | 19.4(270) | 23.7(9) | 23.7(49) | 50.0(1) | |
| It is easier to accept a vaccination certificate than the measures imposed by the pandemic (for ex. partial lockdown, business closures) | | | | | | | | |
| Strongly disagree | 14.7(56) | 10.9(109) | 7.7(80) | 6.6(91) | 2.6(1) | 8.7(18) | 50.0(1) | |
| Disagree | 6.6(25) | 8.7(87) | 10.9(113) | 7.6(105) | 7.9(3) | 8.7(18) | 50.0(1) | |
| Neither agree nor disagree | 22.8(87) | 21.9(219) | 22.4(232) | 17.1(237) | 10.5(4) | 26.1(54) | 0.0(0) | <0.001 |
| Agree | 19.4(74) | 23.9(239) | 24.0(249) | 27.4(380) | 26.3(10) | 23.2(48) | 0.0(0) | |
| Strongly agree | 36.5(139) | 34.7(347) | 35.1(364) | 41.5(576) | 52.6(20) | 33.3(69) | 0.0(0) | |

| | Past SARS-CoV-2 infection | | |
|---|---|---|---|
| | Yes | No | P-value |
| | %(N) | %(N) | |
| **Select the context(s) in which a vaccination certificate should be presented:** | | | |
| If working in a job where others would be at risk (for ex. in nursing homes) | 54.5(420) | 62.1(2,042) | <0.001 |
| If working in a job where the employee is at high risk of infection (for ex. in hospitals) | 51.3(395) | 60.4(1,985) | <0.001 |
| If working in a job where employees have to share the same open workspace | 18.1(139) | 22.3(733) | 0.010 |
| To visit high risk individuals (for ex. in nursing homes or hospitals) | 40.5(312) | 49.3(1,621) | <0.001 |
| To cross national borders | 28.3(218) | 33.1(1,088) | 0.010 |
| To take a plane | 39.1(301) | 45.5(1,494) | 0.001 |
| To be exempt from quarantine when travelling abroad | 47.7(367) | 58.0(1,907) | <0.001 |
| To participate in large gatherings (for ex. concerts, matches etc.) | 31.0(239) | 38.3(1,257) | <0.001 |
| To participate in collective social activities (for ex. cinemas, theater, sports clubs) | 29.7(229) | 36.8(1,210) | <0.001 |
| There is no context where a certificate should be presented | 27.8(214) | 20.1(662) | <0.001 |
| Other contexts | 1.6(12) | 1.3(43) | |
| What is your opinion about the following statements on a scale of 1 (Strongly disagree) to 5 (Strongly agree)? | | | |
| A vaccination certificate should be necessary in certain contexts (for ex. to travel, take care of vulnerable individuals) | | | |
| Strongly disagree | 16.1(124) | 13.0(426) | |
| Disagree | 11.4(88) | 8.1(265) | |
| Neither agree nor disagree | 20.0(154) | 15.9(524) | <0.001 |
| Agree | 22.9(176) | 23.3(765) | |
| Strongly agree | 29.6(228) | 39.7(1,306) | |
| COVID-19 is a trivial disease that does not necessitate a vaccination certificate | | | |
| Strongly disagree | 42.5(327) | 51.6(1,697) | |
| Disagree | 22.1(170) | 20.1(660) | |
| Neither agree nor disagree | 20.3(156) | 16.4(539) | <0.001 |
| Agree | 10.1(78) | 7.4(242) | |
| Strongly agree | 5.1(39) | 4.5(148) | |
| Individuals without a vaccination certificate could be victims of discrimination (for ex. employment opportunities, participating in activities) | | | |
| Strongly disagree | 9.9(76) | 9.3(304) | 0.116 |

| | | | |
|---|---|---|---|
| Disagree | 8.6(66) | 9.6(317) | |
| Neither agree nor disagree | 19.6(151) | 23.4(770) | |
| Agree | 27.8(214) | 26.7(879) | |
| Strongly agree | 34.2(263) | 30.9(1,016) | |
| Individuals without a vaccination certificate risk losing certain rights (for ex. crossing borders) | | | |
| Strongly disagree | 8.3(64) | 7.2(238) | |
| Disagree | 6.2(48) | 7.8(256) | |
| Neither agree nor disagree | 16.9(130) | 18.9(621) | 0.052 |
| Agree | 29.9(230) | 32.1(1,055) | |
| Strongly agree | 38.7(298) | 34.0(1,116) | |
| Personal medical data belongs to the individual and should not be the object of a vaccination certificate | | | |
| Strongly disagree | 19.4(149) | 22.6(741) | |
| Disagree | 14.0(108) | 16.3(537) | |
| Neither agree nor disagree | 19.5(150) | 22.9(752) | <0.001 |
| Agree | 14.7(113) | 12.9(423) | |
| Strongly agree | 32.5(250) | 25.3(833) | |
| It is easier to accept a vaccination certificate than the measures imposed by the pandemic (for ex. partial lockdown, business closures) | | | |
| Strongly disagree | 7.9(61) | 9.0(295) | |
| Disagree | 11.2(86) | 8.1(266) | |
| Neither agree nor disagree | 23.4(180) | 19.9(653) | 0.006 |
| Agree | 23.2(179) | 25.0(821) | |
| Strongly agree | 34.3(264) | 38.1(1,251) | |

| | Did you or will you get vaccinated? | | | |
|---|---|---|---|---|
| | Yes | No | Does not know | P-value |
| | %(N) | %(N) | %(N) | |
| **Select the context(s) in which a vaccination certificate should be presented:** | | | | |
| If working in a job where others would be at risk (for ex. in nursing homes) | 70.0(2,150) | 24.2(136) | 41.6(176) | <0.001 |
| If working in a job where the employee is at high risk of infection (for ex. in hospitals) | 67.6(2,077) | 24.0(135) | 39.7(168) | <0.001 |
| If working in a job where employees have to share the same open workspace | 27.6(849) | 0.9(5) | 4.3(18) | <0.001 |
| To visit high risk individuals (for ex. in nursing homes or hospitals) | 58.4(1,792) | 9.4(53) | 20.8(88) | <0.001 |
| To cross national borders | 40.8(1,254) | 2.7(15) | 8.7(37) | <0.001 |
| To take a plane | 55.6(1,706) | 5.0(28) | 14.4(61) | <0.001 |
| To be exempt from quarantine when travelling abroad | 68.7(2,109) | 11.4(64) | 23.9(101) | <0.001 |
| To participate in large gatherings (for ex. concerts, matches etc.) | 46.1(1,416) | 5.2(29) | 12.1(51) | <0.001 |
| To participate in collective social activities (for ex. cinemas, theater, sports clubs) | 45.0(1,382) | 3.6(20) | 8.7(37) | <0.001 |
| There is no context where a certificate should be presented | 10.9(335) | 66.0(371) | 40.2(170) | <0.001 |
| Other contexts | 1.2(37) | 2.0(11) | 1.7(7) | |
| What is your opinion about the following statements on a scale of 1 (Strongly disagree) to 5 (Strongly agree)? | | | | |
| A vaccination certificate should be necessary in certain contexts (for ex. to travel, take care of vulnerable individuals) | | | | |
| Strongly disagree | 6.3(195) | 45.7(257) | 23.2(98) | |
| Disagree | 5.4(165) | 19.9(112) | 18.0(76) | |
| Neither agree nor disagree | 14.4(442) | 20.6(116) | 28.4(120) | <0.001 |
| Agree | 26.7(819) | 7.5(42) | 18.9(80) | |
| Strongly agree | 47.2(1,450) | 6.2(35) | 11.6(49) | |
| COVID-19 is a trivial disease that does not necessitate a vaccination certificate | | | | |
| Strongly disagree | 61.0(1,873) | 11.9(67) | 19.9(84) | |
| Disagree | 20.0(613) | 21.2(119) | 23.2(98) | |
| Neither agree nor disagree | 10.6(326) | 35.2(198) | 40.4(171) | <0.001 |
| Agree | 4.7(144) | 21.9(123) | 12.5(53) | |
| Strongly agree | 3.7(115) | 9.8(55) | 4.0(17) | |
| Individuals without a vaccination certificate could be victims of discrimination (for ex. employment opportunities, participating in activities) | | | | |
| Strongly disagree | 9.2(282) | 10.7(60) | 9.0(38) | |
| Disagree | 11.0(337) | 4.1(23) | 5.4(23) | |
| Neither agree nor disagree | 26.5(813) | 9.1(51) | 13.5(57) | <0.001 |
| Agree | 28.5(874) | 19.4(109) | 26.0(110) | |
| Strongly agree | 24.9(765) | 56.8(319) | 46.1(195) | |

| | | | | |
|---|---|---|---|---|
| Individuals without a vaccination certificate risk losing certain rights (for ex. crossing borders) | | | | |
| Strongly disagree | 6.6(203) | 11.0(62) | 8.7(37) | |
| Disagree | 8.2(251) | 5.2(29) | 5.7(24) | |
| Neither agree nor disagree | 20.7(636) | 8.4(47) | 16.1(68) | <0.001 |
| Agree | 34.7(1,066) | 20.3(114) | 24.8(105) | |
| Strongly agree | 29.8(915) | 55.2(310) | 44.7(189) | |
| Personal medical data belongs to the individual and should not be the object of a vaccination certificate | | | | |
| Strongly disagree | 27.2(834) | 6.4(36) | 4.7(20) | |
| Disagree | 19.3(592) | 3.4(19) | 8.0(34) | |
| Neither agree nor disagree | 24.5(751) | 10.9(61) | 21.3(90) | <0.001 |
| Agree | 12.5(383) | 14.8(83) | 16.5(70) | |
| Strongly agree | 16.6(511) | 64.6(363) | 49.4(209) | |
| It is easier to accept a vaccination certificate than the measures imposed by the pandemic (for ex. partial lockdown, business closures) | | | | |
| Strongly disagree | 4.8(146) | 26.0(146) | 15.1(64) | |
| Disagree | 4.9(152) | 21.0(118) | 19.4(82) | |
| Neither agree nor disagree | 16.9(519) | 32.2(181) | 31.4(133) | <0.001 |
| Agree | 27.5(843) | 12.6(71) | 20.3(86) | |
| Strongly agree | 45.9(1,411) | 8.2(46) | 13.7(58) | |

| | Is vaccination an important step to surmount the current COVID-19 pandemic? | | |
|---|---|---|---|
| | Yes or rather yes | No or rather no | P-value |
| | %(N) | %(N) | |
| **Select the context(s) in which a vaccination certificate should be presented:** | | | |
| If working in a job where others would be at risk (for ex. in nursing homes) | 64.5(2,414) | 15.3(48) | <0.001 |
| If working in a job where the employee is at high risk of infection (for ex. in hospitals) | 62.2(2,327) | 16.9(53) | <0.001 |
| If working in a job where employees have to share the same open workspace | 23.2(870) | 0.6(2) | <0.001 |
| To visit high risk individuals (for ex. in nursing homes or hospitals) | 51.2(1,915) | 5.8(18) | <0.001 |
| To cross national borders | 34.7(1,300) | 1.9(6) | <0.001 |
| To take a plane | 47.7(1,786) | 2.9(9) | <0.001 |
| To be exempt from quarantine when travelling abroad | 60.1(2,251) | 7.3(23) | <0.001 |
| To participate in large gatherings (for ex. concerts, matches etc.) | 39.7(1,486) | 3.2(10) | <0.001 |
| To participate in collective social activities (for ex. cinemas, theater, sports clubs) | 38.3(1,433) | 1.9(6) | <0.001 |
| There is no context where a certificate should be presented | 17.0(636) | 76.7(240) | <0.001 |
| Other contexts | 1.4(53) | 0.6(2) | |
| What is your opinion about the following statements on a scale of 1 (Strongly disagree) to 5 (Strongly agree)? | | | |
| A vaccination certificate should be necessary in certain contexts (for ex. to travel, take care of vulnerable individuals) | | | |
| Strongly disagree | 9.9(372) | 56.9(178) | |
| Disagree | 7.9(297) | 17.9(56) | |
| Neither agree nor disagree | 16.8(628) | 16.0(50) | <0.001 |
| Agree | 24.8(930) | 3.5(11) | |
| Strongly agree | 40.5(1,516) | 5.8(18) | |
| COVID-19 is a trivial disease that does not necessitate a vaccination certificate | | | |
| Strongly disagree | 53.4(1,997) | 8.6(27) | |
| Disagree | 20.8(778) | 16.6(52) | |
| Neither agree nor disagree | 15.8(592) | 32.9(103) | <0.001 |
| Agree | 6.4(241) | 25.2(79) | |
| Strongly agree | 3.6(135) | 16.6(52) | |
| Individuals without a vaccination certificate could be victims of discrimination (for ex. employment opportunities, participating in activities) | | | |
| Strongly disagree | 9.1(342) | 12.1(38) | <0.001 |

| | | | |
|---|---|---|---|
| Disagree | 9.9(371) | 3.8(12) | |
| Neither agree nor disagree | 23.8(890) | 9.9(31) | |
| Agree | 27.7(1,036) | 18.2(57) | |
| Strongly agree | 29.5(1,104) | 55.9(175) | |
| Individuals without a vaccination certificate risk losing certain rights (for ex. crossing borders) | | | |
| Strongly disagree | 7.0(262) | 12.8(40) | |
| Disagree | 7.7(289) | 4.8(15) | |
| Neither agree nor disagree | 19.1(714) | 11.8(37) | <0.001 |
| Agree | 32.9(1,233) | 16.6(52) | |
| Strongly agree | 33.3(1,245) | 54.0(169) | |
| Personal medical data belongs to the individual and should not be the object of a vaccination certificate | | | |
| Strongly disagree | 23.5(880) | 3.2(10) | |
| Disagree | 17.1(639) | 1.9(6) | |
| Neither agree nor disagree | 23.4(874) | 8.9(28) | <0.001 |
| Agree | 13.3(499) | 11.8(37) | |
| Strongly agree | 22.7(851) | 74.1(232) | |
| It is easier to accept a vaccination certificate than the measures imposed by the pandemic (for ex. partial lockdown, business closures) | | | |
| Strongly disagree | 6.6(248) | 34.5(108) | |
| Disagree | 7.7(288) | 20.4(64) | |
| Neither agree nor disagree | 19.7(737) | 30.7(96) | <0.001 |
| Agree | 26.0(974) | 8.3(26) | |
| Strongly agree | 40.0(1,496) | 6.1(19) | |
